# Shared Genomic Architectures of COVID-19 and Antisocial Behavior

**DOI:** 10.1101/2021.10.18.21265145

**Authors:** Charleen D. Adams, Jorim J. Tielbeek, Brian B. Boutwell

**Affiliations:** Department of Environmental Health, Program in Molecular and Integrative Physiological Sciences, Harvard T.H. Chan School of Public Health; Department of Complex Trait Genetics, Center for Neurogenomics and Cognitive Research, Amsterdam Neuroscience, Vrije Universiteit Amsterdam, Amsterdam, Netherlands; School of Applied Sciences, The University of Mississippi; John D. Bower School of Population Health, University of Mississippi Medical Center

## Abstract

Little is known about the genetics of norm violation and aggression (ASB) in relation to coronavirus disease 2019 (COVID-19). To investigate this, we used summary statistics from genome-wide association studies and linkage disequilibrium score regression to calculate a matrix of genetic correlations (*r*_*gs*_) for ASB, COVID-19, and various health and behavioral traits. After false-discovery rate correction, ASB was genetically correlated with COVID-19 (*r*_*g*_ = 0.51; *P* = 1.54E-02) and 19 other traits. ASB and COVID-19 were both positively genetically correlated with having a noisy workplace, doing heavy manual labor, chronic obstructive pulmonary disease, and genitourinary diseases. ASB and COVID-19 were both inversely genetically correlated with average income, education years, healthspan, verbal reasoning, lifespan, cheese intake, and being breastfed as a baby. But keep in mind that *r*_*gs*_ are not necessarily causal. And, if causal, their prevailing directions of effect (which causes which) are indiscernible from *r*_*gs*_ alone. Moreover, the SNP-heritability 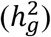 estimates for both measures of COVID-19 were very low, restricting the overlap of genetic variance in absolute terms between the two traits. Nonetheless, our findings suggest that those with antisocial tendencies possibly have a higher risk of exposure to severe acute respiratory syndrome coronavirus 2 (SARS-CoV-2) than those without antisocial tendencies. This may have been especially true early in the pandemic before vaccines against SARS-CoV-2 were available and before the emergence of the highly transmissible Omicron variant.

## Introduction

Antisocial behavior (ASB)— including aggression, rule-breaking, delinquency, and violence— are harmful to society. ASB creates a long wake of monetary, social, and emotional disturbances for countries, communities, and individuals^1,2^. Especially troublesome are the possible effects during pandemics. For instance, ASB may abet pandemic spread. Those engaged in overt ASB seem to adhere less to coronavirus disease 2019 (COVID-19) containment measures^3–5^. Similarly, individuals scoring higher on less obvious indicators of antisociality (i.e., low acceptance of moral rules, higher levels of psychopathy, “pre-pandemic legal cynicism, low shame/guilt, low self-control, engagement in delinquent behaviors, and association with delinquent peers”) have shown evidence of disregarding public-health guidelines^3,4,6^. This warrants further investigation into the possible connections between ASB and exposure to severe acute respiratory syndrome coronavirus 2 (SARS-CoV-2), the virus that causes COVID-19.

Complicating causal inference concerning the links between of ASB and pandemic-relevant outcomes is that about half of the variance in ASB and, to varying degrees, associated traits, is heritable^7–9^. This matters because the extent to which ASB and other traits share genetic architecture could influence the likelihood of genetic confounding in observational studies. Broadly addressing this problem is a nascent area of research that uses genome-wide association (GWA) studies of ASB and various health and behavioral traits to calculate genetic correlations (*r*_*gs*_)^10^. These studies have revealed *r*_*gs*_ between ASB and most psychiatric, psychological, reproductive, cognitive, and addictive traits^11,12^. In addition, those prone to antisocial, violent, and criminal behaviors are disproportionately and profoundly unhealthy^13,14^. A strongly negative genetic correlation (*r*_*g*_ = -0.55) between ASB and self-reported health has been reported^11^. In contrast, a comprehensive study found no significant *r*_*gs*_ between ASB and 669 health, physiological, and well-being measures after accounting for multiple testing^15^. Thus, much remains to be discovered regarding shared etiology between ASB and various aspects of health, including COVID-19.

## Methods and materials

We characterized the shared polygenic nature of ASB, COVID-19, and various health and behavioral traits using summary statistics from GWA studies and linkage disequilibrium score regression (LDSC; software available at http://www.github.com/bulik/ldsc)^16^. We calculated a matrix of *r*_*gs*_. Of note is that correlation, even when genetic, is not necessarily causation. While our study can point to shared genetic architecture between traits, the reader should be cautious about assuming that the *r*_*gs*_ are causal. **Table 1** contains details about the GWA studies we used and where interested researchers can access them. Nineteen traits were chosen for novelty (having not been previously reported as either null or significantly correlated with ASB). The novel traits include: average income (before taxes); healthspan (i.e., living free from congestive heart failure, myocardial infarction, chronic obstructive pulmonary disease [COPD], stroke, dementia, diabetes, cancer, and death; coded as a protective ratio); parental lifespan (hereafter “lifespan”; coded as a protective ratio); word interpolation (hereafter “verbal reasoning”); having been breastfed as baby; cheese intake; self-reported happiness; having had COVID-19 (data from two GWA studies); doing heavy manual labor; having a noisy workplace; Townsend Deprivation Index (an area- and census-based measure of deprivation, where a higher score indicates more deprivation); having gastrointestinal diseases; having COPD; having genitourinary diseases; playing computer games; having been a violent-crime victim; risk tolerance, and witnessing a sudden violent death. Four traits (education years; seen doctor for nerves, anxiety, tension, or depression; neuroticism; and Parkinson’s disease) were chosen as replicates of previously reported findings.

**Table 1.**
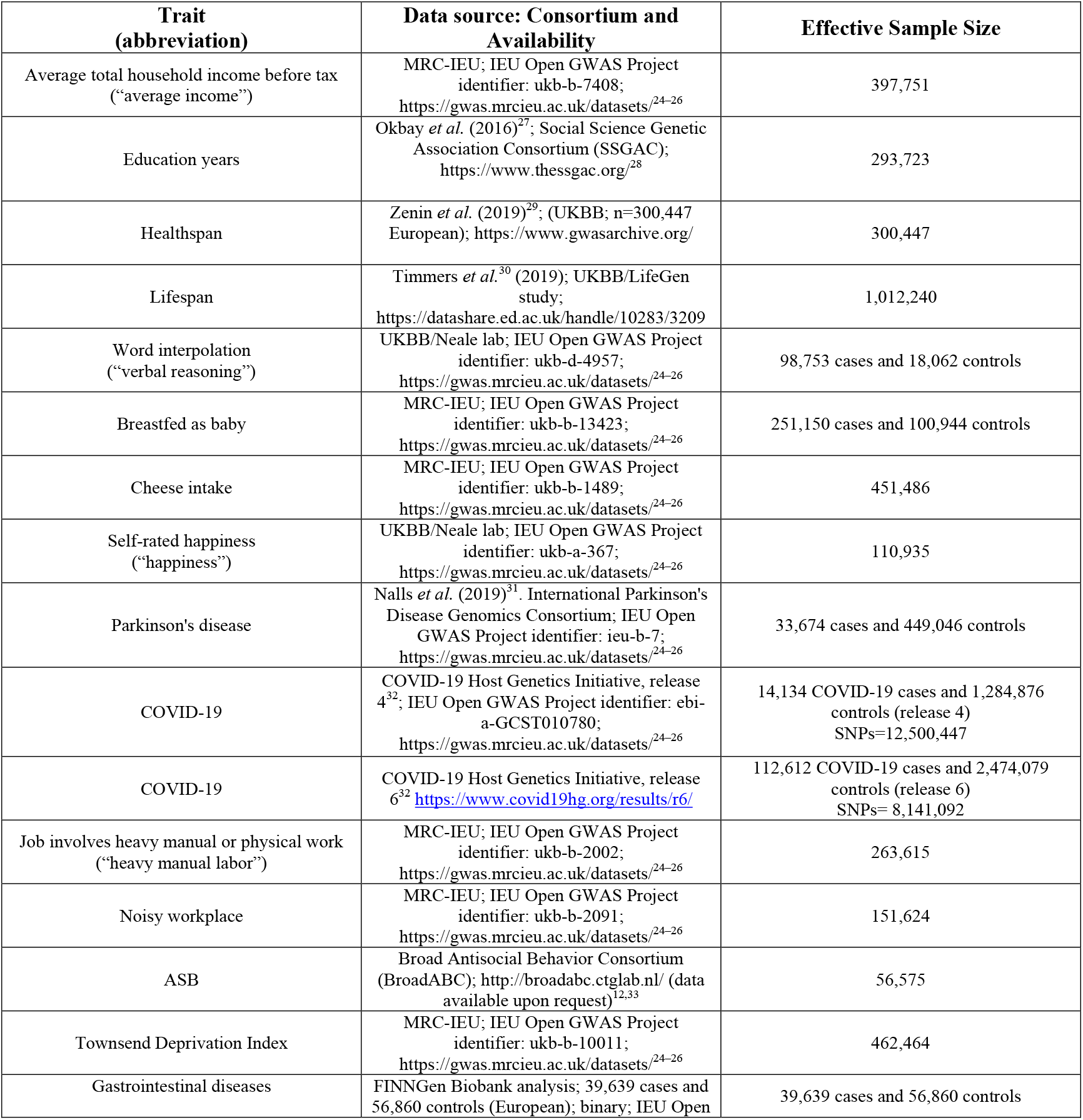
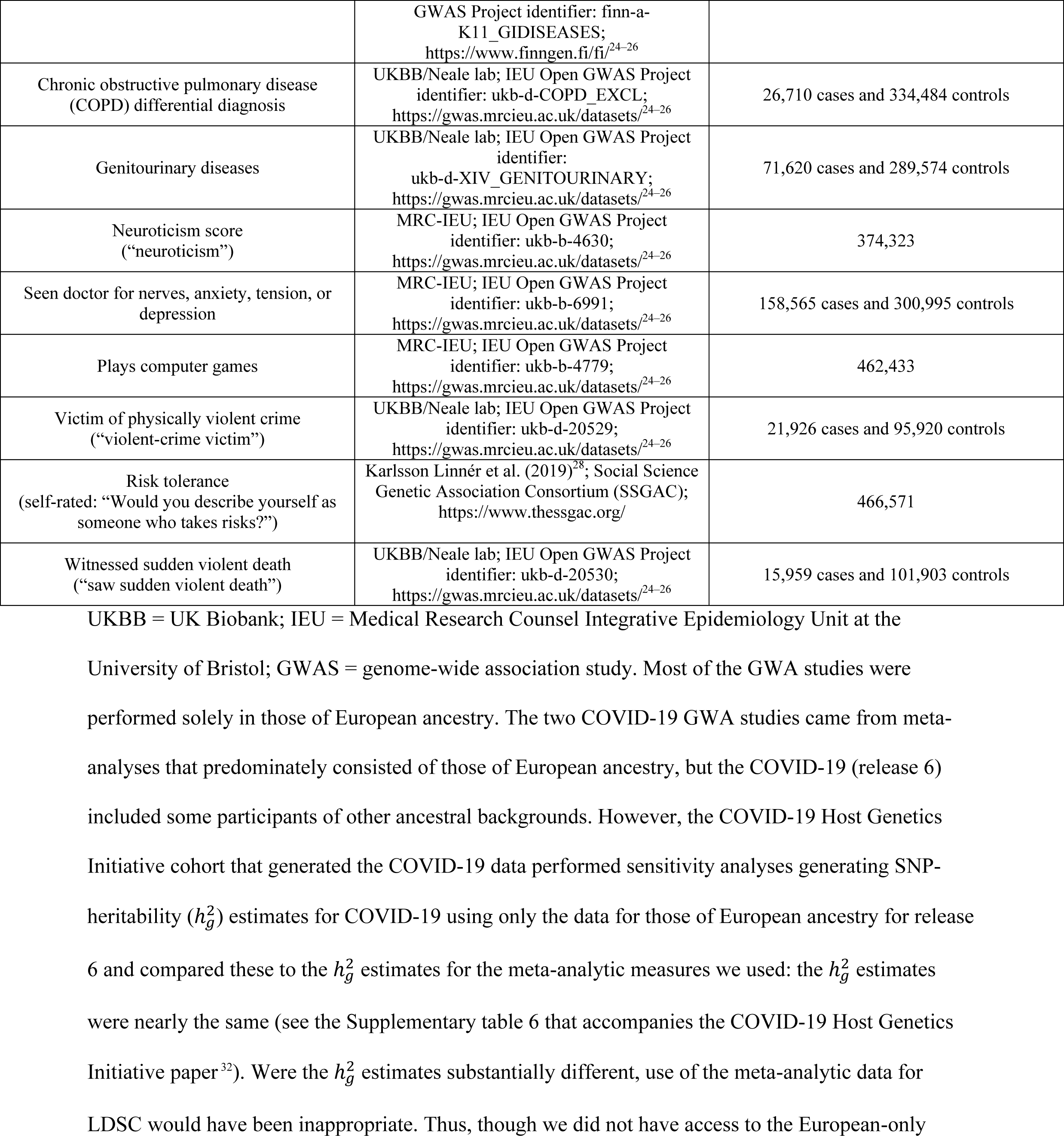
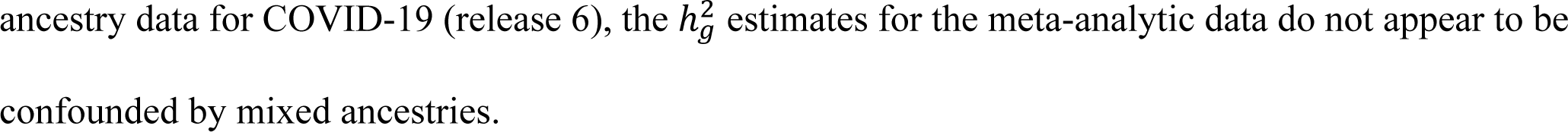
GWA study data sources.

## Results

### ASB

After false-discovery rate (FDR)-correction (*P* < 0.05), ASB was positively genetically correlated with COVID-19 (release 4): *r*_*g*_ = 0.51; *P* = 1.54E-02. The *r*_*g*_ was also positive between ASB and COVID-19 (release 6) with marginal significance *prior* to FDR-correction but not after: *r*_*gs*_ = 0.35; *P* = 3.83E-02 (FDR-corrected *P* = 5.21E-02). The remaining (FDR-significant) *r*_*gs*_ between ASB and health and behavioral traits that were positive are as follows:

1. Townsend Deprivation Index (*r*_*g*_ = 0.70)
2. Noisy workplace (*r*_*g*_ = 0.63)
3. Heavy manual labor (*r*_*g*_ = 0.58)
4. COPD (*r*_*g*_ = 0.51)
5. Risk tolerance (*r*_*g*_ = 0.50)
6. Gastrointestinal diseases (*r*_*g*_ = 0.46)
7. Seen a doctor for nerves, anxiety, tension, or depression (*r*_*g*_ = 0.42)
8. Seen a sudden violent death (*r*_*g*_ = 0.42)
9. Genitourinary diseases (*r*_*g*_ = 0.38)
10. Being a violent-crime victim (*r*_*g*_ = 0.36)
11. Neuroticism (*r*_*g*_ = 0.29)
12. Playing computer games (*r*_*g*_ = 0.15)

ASB was negatively genetically correlated with seven traits (after FDR-correction):

1. Average income (*r*_*g*_ = -0.54)
2. Education years (*r*_*g*_ = -0.48)
3. Healthspan (*r*_*g*_ = -0.47)
4. Verbal reasoning (*r*_*g*_ = -0.44)
5. Lifespan (*r*_*g*_ = -0.33)
6. Cheese intake (*r*_*g*_ = -0.28)
7. Breastfed as baby (*r*_*g*_ = -0.24)

The *r*_*gs*_ for ASB and the health and behavioral traits are displayed in a forest plot in **Figure 1** and presented in **Table 2** along with confidence intervals and SNP-heritability 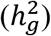 estimates. See the Supplement for the results for all the *r*_*gs*_ in the matrix (Supplementary Table 1), including the *p*-values before and after FDR-correction (Supplementary Table 2).

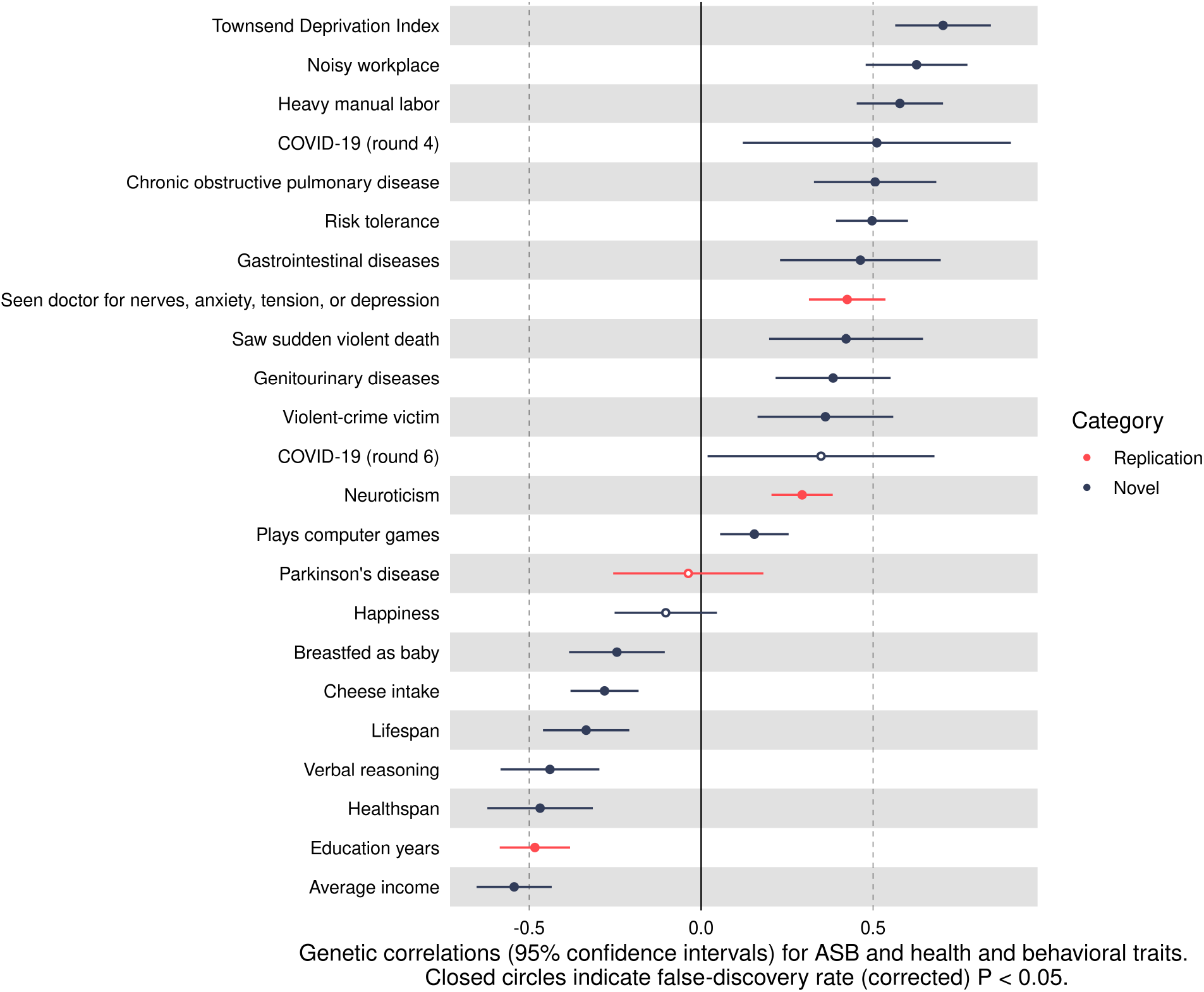

**Table 2.** Genetic correlations (*r*_*gs*_) between ASB and health and behavioral traits.

| Trait 1 | Trait 2 | $r_g$ | Lower<br>95% CI<br>for $r_g$ | Upper<br>95% CI<br>for $r_g$ | FDR $P$-<br>value | $h_g^2$ for<br>trait 2 |
| --- | --- | --- | --- | --- | --- | --- |
| ASB | Average income | -0.54 | -0.65 | -0.43 | 9.88E-22 | 0.07 |
| ASB | Education years | -0.48 | -0.59 | -0.38 | 9.76E-20 | 0.12 |
| ASB | Healthspan | -0.47 | -0.62 | -0.31 | 5.97E-09 | 0.03 |
| ASB | Verbal reasoning | -0.44 | -0.58 | -0.30 | 5.62E-09 | 0.08 |
| ASB | Lifespan | -0.33 | -0.46 | -0.21 | 4.20E-07 | 0.02 |
| ASB | Cheese intake | -0.28 | -0.38 | -0.18 | 6.97E-08 | 0.07 |
| ASB | Breastfed as baby | -0.24 | -0.38 | -0.11 | 9.60E-04 | 0.03 |
| ASB | Happiness | -0.10 | -0.25 | 0.05 | 2.22E-01 | 0.06 |
| ASB | Parkinson's disease | -0.04 | -0.26 | 0.18 | 7.77E-01 | 0.02 |
| ASB | Plays computer games | 0.15 | 0.06 | 0.25 | 3.60E-03 | 0.07 |
| ASB | Neuroticism | 0.29 | 0.20 | 0.38 | 3.05E-10 | 0.11 |
| ASB | COVID-19 (release 6) | 0.35 | 0.02 | 0.68 | 5.21E-02 | 0.001 |
| ASB | Violent-crime victim | 0.36 | 0.16 | 0.56 | 5.82E-04 | 0.03 |
| ASB | Genitourinary diseases | 0.38 | 0.22 | 0.55 | 1.45E-05 | 0.02 |
| ASB | Saw sudden violent death | 0.42 | 0.20 | 0.65 | 3.95E-04 | 0.02 |
| ASB | Seen doctor for nerves, anxiety, tension, or depression | 0.42 | 0.31 | 0.54 | 2.36E-13 | 0.06 |
| ASB | Gastrointestinal diseases | 0.46 | 0.23 | 0.70 | 1.89E-04 | 0.04 |
| ASB | Risk tolerance | 0.50 | 0.39 | 0.60 | 6.34E-20 | 0.02 |
| ASB | COPD | 0.51 | 0.33 | 0.68 | 6.45E-08 | 0.01 |
| ASB | COVID-19 (release 4) | 0.51 | 0.12 | 0.90 | 1.54E-02 | 0.001 |
| ASB | Heavy manual labor | 0.58 | 0.45 | 0.70 | 8.31E-19 | 0.08 |
| ASB | Noisy workplace | 0.63 | 0.48 | 0.77 | 3.99E-16 | 0.06 |
| ASB | Townsend Deprivation Index | 0.70 | 0.56 | 0.84 | 2.25E-22 | 0.03 |

ASB=antisocial behavior; $r_g$ = genetic correlation; FDR = false-discovery rate (corrected) $P$ -value; $h_g^2$
= SNP-heritability.

### COVID-19

Due to the positive *r*_*g*_ between COVID-19 and ASB, we highlight the FDR-significant *r*_*gs*_ between COVID-19 and non-ASB traits. COVID-19 was positively genetically correlated with the following:

1. COPD (*r*_*g*_ = 0.40) -- COVID-19 (release 6)
2. COPD (*r*_*g*_ = 0.33) -- COVID-19 (release 4)
3. Heavy manual labor (*r*_*g*_ = 0.38) -- COVID-19 (release 6)
4. Heavy manual labor (*r*_*g*_ = 0.20) -- COVID-19 (release 4)
5. Genitourinary diseases (*r*_*g*_ = 0.32) -- COVID-19 (release 6)
6. Noisy workplace (*r*_*g*_ = 0.28) -- COVID-19 (release 6)
7. Noisy workplace (*r*_*g*_ = 0.26) -- COVID-19 (release 4)

COVID-19 was negatively genetically correlated with the following:

1. Cheese intake (*r*_*g*_ = -0.39) -- COVID-19 (release 6)
2. Cheese intake (*r*_*g*_ = -0.36) -- COVID-19 (release 4)
3. Education years (*r*_*g*_ = -0.46) -- COVID-19 (release 6)
4. Education years (*r*_*g*_ = -0.32) -- COVID-19 (release 4)
5. Verbal reasoning (*r*_*g*_ = -0.49) -- COVID-19 (release 6)
6. Verbal reasoning (*r*_*g*_ = -0.28) -- COVID-19 (release 4)
7. Healthspan (*r*_*g*_ = -0.41) -- COVID-19 (release 6)
8. Healthspan (*r*_*g*_ = -0.25) -- COVID-19 (release 4)
9. Breastfed as baby (*r*_*g*_ = -0.24) -- COVID-19 (release 6)
10. Lifespan (*r*_*g*_ = -0.30) -- COVID-19 (release 6)
11. Average income (*r*_*g*_ = -0.21) -- COVID-19 (release 6)

Notably, ASB and COVID-19 were both positively genetically correlated with having a noisy workplace, doing heavy manual labor, COPD, and genitourinary diseases. They were both inversely genetically correlated with average income, education years, healthspan, verbal reasoning, lifespan, cheese intake, and being breastfed as a baby. The *r*_*gs*_ between COVID-19 and the non-ASB traits are presented in **Figure 2** and **Table 3** along with confidence intervals and 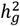 estimates.

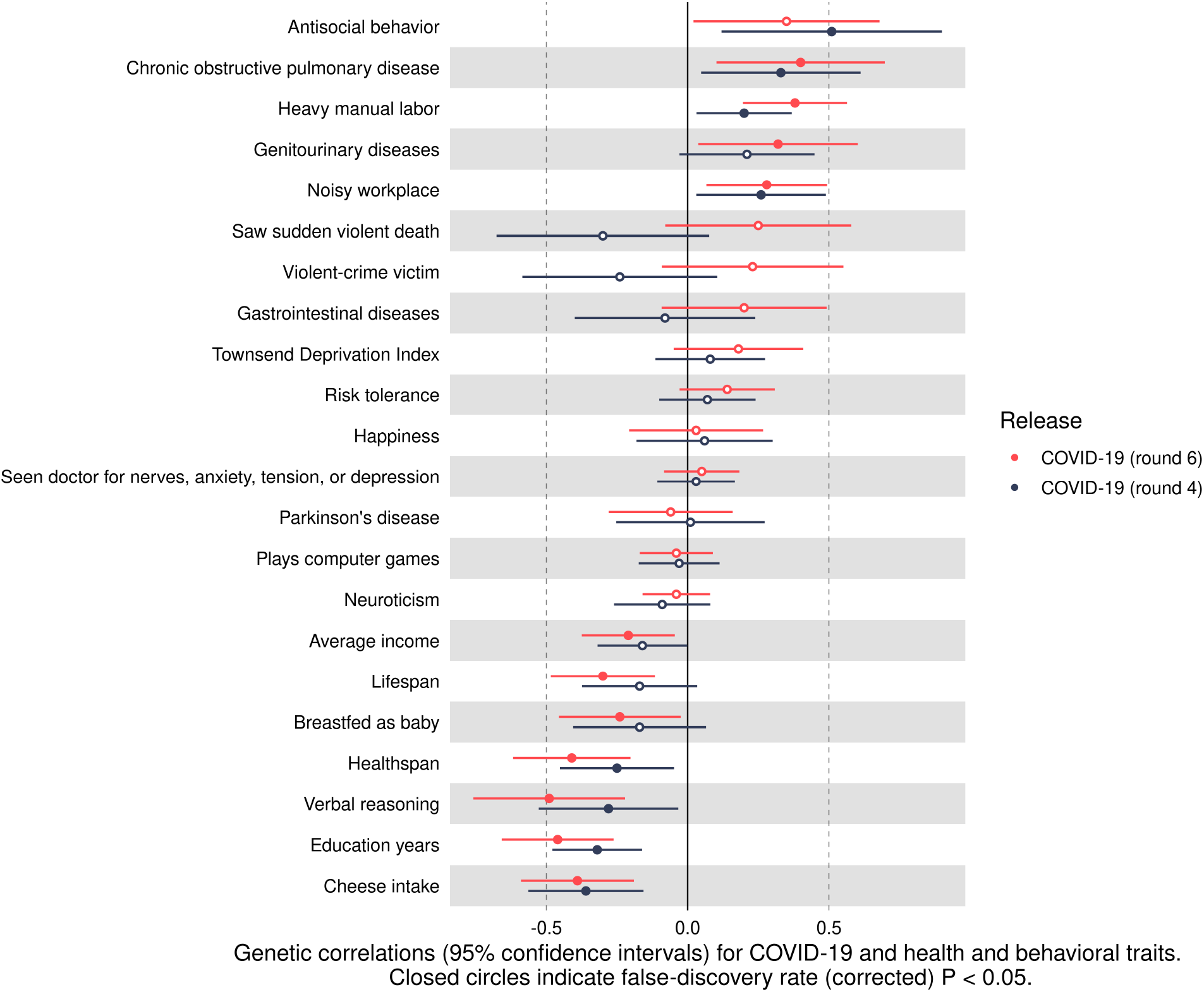

**Table 3.** Genetic correlations (*r*_*gs*_) between COVID-19 and health and behavioral traits.

| Trait 1 | Trait 2 | $r_g$ | Lower<br>95% CI<br>for $r_g$ | Upper<br>95% CI<br>for $r_g$ | FDR $P$-<br>value | $h_g^2$ for<br>trait 2 |
| --- | --- | --- | --- | --- | --- | --- |
| COVID-19 (release 6) | Chronic obstructive pulmonary disease | 0.40 | 0.11 | 0.70 | 1.17E-02 | 0.02 |
| COVID-19 (release 6) | Heavy manual labor | 0.38 | 0.19 | 0.56 | 1.25E-04 | 0.08 |
| COVID-19 (release 4) | Chronic obstructive pulmonary disease | 0.33 | 0.04 | 0.61 | 3.44E-02 | 0.01 |
| COVID-19 (release 6) | Genitourinary diseases | 0.32 | 0.04 | 0.60 | 3.69E-02 | 0.02 |
| COVID-19 (release 6) | Noisy workplace | 0.28 | 0.07 | 0.50 | 1.45E-02 | 0.06 |
| COVID-19 (release 4) | Noisy workplace | 0.26 | 0.03 | 0.49 | 3.69E-02 | 0.06 |
| COVID-19 (release 6) | Saw sudden violent death | 0.25 | -0.08 | 0.58 | 1.70E-01 | 0.03 |
| COVID-19 (release 6) | Violent-crime victim | 0.23 | -0.09 | 0.55 | 1.97E-01 | 0.03 |
| COVID-19 (release 4) | Genitourinary diseases | 0.21 | -0.03 | 0.45 | 1.17E-01 | 0.02 |
| COVID-19 (release 4) | Heavy manual labor | 0.20 | 0.03 | 0.37 | 2.65E-02 | 0.08 |
| COVID-19 (release 6) | Gastrointestinal diseases | 0.20 | -0.10 | 0.49 | 2.36E-01 | 0.05 |
| COVID-19 (release 6) | Townsend Deprivation Index | 0.18 | -0.05 | 0.41 | 1.66E-01 | 0.03 |
| COVID-19 (release 6) | Risk tolerance | 0.14 | -0.03 | 0.31 | 1.31E-01 | 0.02 |
| COVID-19 (release 4) | Townsend Deprivation Index | 0.08 | -0.11 | 0.27 | 4.81E-01 | 0.03 |
| COVID-19 (release 4) | Risk tolerance | 0.07 | -0.10 | 0.24 | 4.97E-01 | 0.02 |
| COVID-19 (release 4) | Happiness | 0.06 | -0.19 | 0.30 | 7.07E-01 | 0.06 |
| COVID-19 (release 6) | Seen doctor for nerves, anxiety, tension, or depression | 0.05 | -0.09 | 0.18 | 5.40E-01 | 0.06 |
| COVID-19 (release 4) | Seen doctor for nerves, anxiety, tension, or depression | 0.03 | -0.11 | 0.17 | 7.22E-01 | 0.06 |
| COVID-19 (release 6) | Happiness | 0.03 | -0.21 | 0.27 | 8.38E-01 | 0.06 |
| COVID-19 (release 4) | Parkinson's disease | 0.01 | -0.26 | 0.27 | 9.59E-01 | 0.02 |
| COVID-19 (release 4) | Plays computer games | -0.03 | -0.17 | 0.11 | 7.19E-01 | 0.07 |
| COVID-19 (release 6) | Plays computer games | -0.04 | -0.17 | 0.09 | 6.23E-01 | 0.08 |
| COVID-19 (release 6) | Neuroticism | -0.04 | -0.16 | 0.08 | 5.25E-01 | 0.11 |
| COVID-19 (release 6) | Parkinson's disease | -0.06 | -0.28 | 0.16 | 6.61E-01 | 0.02 |
| COVID-19 (release 4) | Gastrointestinal diseases | -0.08 | -0.39 | 0.24 | 7.06E-01 | 0.04 |
| COVID-19 (release 4) | Neuroticism | -0.09 | -0.26 | 0.08 | 3.68E-01 | 0.11 |
| COVID-19 (release 4) | Average income | -0.16 | -0.32 | 0.00 | 6.02E-02 | 0.07 |
| COVID-19 (release 4) | Lifespan | -0.17 | -0.37 | 0.04 | 1.47E-01 | 0.02 |
| COVID-19 (release 4) | Breastfed as baby | -0.17 | -0.41 | 0.06 | 1.93E-01 | 0.03 |
| COVID-19 (release 6) | Average income | -0.21 | -0.37 | -0.04 | 2.06E-02 | 0.07 |
| COVID-19 (release 4) | Violent-crime victim | -0.24 | -0.58 | 0.11 | 2.24E-01 | 0.03 |
| COVID-19 (release 6) | Breastfed as baby | -0.24 | -0.46 | -0.03 | 3.69E-02 | 0.02 |
| COVID-19 (release 4) | Healthspan | -0.25 | -0.45 | -0.04 | 2.53E-02 | 0.03 |
| COVID-19 (release 4) | Verbal reasoning | -0.28 | -0.52 | -0.03 | 3.79E-02 | 0.08 |
| COVID-19 (release 6) | Lifespan | -0.30 | -0.48 | -0.11 | 2.73E-03 | 0.02 |
| COVID-19 (release 4) | Saw sudden violent death | -0.30 | -0.68 | 0.07 | 1.50E-01 | 0.03 |
| COVID-19 (release 4) | Education years | -0.32 | -0.48 | -0.16 | 1.39E-04 | 0.13 |
| COVID-19 (release 4) | Cheese intake | -0.36 | -0.56 | -0.16 | 9.65E-04 | 0.07 |
| COVID-19 (release 6) | Cheese intake | -0.39 | -0.59 | -0.20 | 1.88E-04 | 0.07 |
| COVID-19 (release 6) | Healthspan | -0.41 | -0.62 | -0.20 | 2.23E-04 | 0.03 |
| COVID-19 (release 6) | Education years | -0.46 | -0.66 | -0.26 | 1.09E-05 | 0.13 |
| COVID-19 (release 6) | Verbal reasoning | -0.49 | -0.76 | -0.22 | 6.28E-04 | 0.09 |

$r_g$ = genetic correlation; FDR = false-discovery rate (corrected) $P$ -value; $h_g^2$ = SNP-heritability.

## Discussion

In support of prior observational findings by O’Connell *et al*. (2021)^5^, Carvalho and Machado (2020)^3^, Miguel *et al*. (2021)^4^, and Nivette *et al*. (2020)^6^, the positive *r*_*g*_ between ASB and COVID-19 suggests that those with antisocial tendencies are more likely to be exposed to SARS-CoV-2 than those who do not engage in ASB. Although ASB is generally associated with impulsive and risk-taking proclivities, the *r*_*g*_ between COVID-19 and risk tolerance was null in our study, a result that argues against a propensity for risk-taking behavior underlying the link between ASB and exposure to SARS-CoV-2. The totality of our data instead suggests that a broad architecture of factors predispose some to both ASB and COVID-19. Traits, for example, that are positively genetically correlated with both ASB and COVID-19—having a noisy workplace, doing heavy manual labor, and having COPD—are also strongly inversely genetically correlated with education years, verbal reasoning, and average income.

We observed positive *r*_*gs*_ between ASB and the psychiatric and violence-related traits we measured. But none of these traits were genetically correlated with COVID-19. That they were not comports with a meta-analytic review of mood disorders and risk for COVID-19 in 91 million individuals^17^. Namely, Ceban *et al*. (2021) found no association between pre-existing mood disorders and COVID-19^17^. Thus, the link between ASB and COVID-19 is unlikely to be due to those engaging in ASB having comorbid mood disorders.

We note that the strength of the *r*_*g*_ for ASB and COVID-19 dropped from 0.51 (release 4) to 0.35 (release 6). Earlier GWA study releases by the COVID-19 Host Genetics Initiative capture data from earlier timepoints in the pandemic—release 4 being earlier (October 20, 2020) than release 6 (June 15, 2021). This may be important since release 4 occurred before vaccines against SARS-CoV-2 were available, and by June 15, 2021, 47% of those eligible for vaccination had completed an initial protocol for full vaccination in the U.S.^18^. Also, both releases 4 and 6 occurred prior to the appearance of the more transmissible Omicron (B.1.1.529) variant, which most on the planet are expected to encounter eventually^19,20^. Thus, our results seem to reflect an increased risk of exposure to SARS-CoV-2 early in the pandemic for those prone to ASB. If those with antisocial tendencies disproportionately refuse vaccination against SARS-CoV-2, however, the impact of ASB over time may have shifted from who gets exposed to SARS-CoV-2 to who gets severe disease.

Our study has limitations, which must also be considered. First is that the 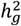 estimates for both measures of COVID-19, while >0, were very small. This indicates that, based on the GWA results, SNPs can only explain a very small proportion of the individual differences in risk for COVID-19. Hence, although the *r*_*gs*_ between ASB and COVID-19 were considerable, in absolute terms the genetic variance that is overlapping between the two traits is low. Second is that *r*_*gs*_, while robust against most environmental confounders, can still suffer from genetic sources of confounding (i.e., even with *r*_*gs*_, as we mentioned above, correlation is not necessarily causation). To illustrate, it seems unintuitive and unlikely that not being breastfed as a baby and eating less cheese cause ASB. One should, for somewhat obvious reasons, be similarly cautioned against the conclusion that being breastfed as a baby and eating more cheese protect against COVID-19, despite the apparent *r*_*gs*_. Indeed, we chose these dietary traits to communicate the point that the shared genetic architecture that these have with education years, verbal reasoning, and average income seem the more plausibly causal phenomenon. Third, supposing that some of the *r*_*gs*_ represent causal linkages in some way, we nonetheless cannot determine the direction of causality with *r*_*gs*_ alone. For much of the discussion above, we tacitly presumed plausible directions of effect (e.g., ASB causing exposure to SARS-CoV-2 and, thus, COVID-19 versus COVID-19 causing ASB). But with all the traits in our matrix, the prevailing direction of effect could be the opposite and/or some level of bi-directional causation may exist^16,21–23^. And, as alluded to by “shared genetic architecture,” the correlated traits could be tagging a latent causal factor. These uncertainties are avenues for future research. Future studies could use either latent causal variable (LCV)^22^ models to infer causality between traits or perform bi-directional MR, an instrumental variables technique, for which both directions of effect are probed. Regarding MR, few genome-wide significant signals have been found for ASB, and using SNPs weakly associated with ASB as instrumental variables would violate the assumptions necessary to perform MR. But assuming SNPs strongly associated with ASB are eventually found, bi-directional MR can be used to decipher the prevailing directions of effect between ASB and traits with which it’s associated. A fourth limitation is that our findings are limited to those of European ancestry. The limitations notwithstanding, *r*_*gs*_ obtained from LDSC are not affected by sample overlap (i.e., participants being in both GWA studies for which the *r*_*gs*_ were calculated)^16^. This is a strength of study, which enabled us to capitalize on the power of large, population-based cohorts and publicly available GWA studies to probe timely questions. Finally, working to understand the etiology of ASB gets us closer to thinking about strategies to provide relief to a large part of the global population—both those engaged in ASB and those devastated by it.

## Data Availability

All data produced in the study are available in the Supplementary Appendix.

## Acknowledgements

We thank the consortia that made their GWA studies public.

## Conflicts of Interest

The authors declare no conflicts of interest.

## Notes

### Competing Interest Statement

The authors have declared no competing interest.

### Funding Statement

This study did not receive any funding.

### Author Declarations

The data are openly available at the following locations: 1.Average total household income before tax: MRC-IEU; IEU Open GWAS Project identifier: ukb-b-7408; https://gwas.mrcieu.ac.uk/datasets/. 2.Education years: Okbay et al. (2016); Social Science Genetic Association Consortium (SSGAC); https://www.thessgac.org/ 3.Healthspan: Zenin et al. (2019); (UKBB; n=300,447 European); https://www.gwasarchive.org/ 4.Lifespan: Timmers et al. (2019); UKBB/LifeGen study; https://datashare.ed.ac.uk/handle/10283/3209 5.Word interpolation: UKBB/Neale lab; IEU Open GWAS Project identifier: ukb-d-4957; https://gwas.mrcieu.ac.uk/datasets/ 6.Breastfed as baby: MRC-IEU; IEU Open GWAS Project identifier: ukb-b-13423; https://gwas.mrcieu.ac.uk/datasets/ 7.Cheese intake: MRC-IEU; IEU Open GWAS Project identifier: ukb-b-1489; https://gwas.mrcieu.ac.uk/datasets/ 8.Happiness: UKBB/Neale lab; IEU Open GWAS Project identifier: ukb-a-367; https://gwas.mrcieu.ac.uk/datasets/ 9.Parkinson's disease: Nalls et al. (2019). International Parkinson's Disease Genomics Consortium; IEU Open GWAS Project identifier: ieu-b-7; https://gwas.mrcieu.ac.uk/datasets/ 10.Covid-19 (round 4): Covid-19 Host Genetics Initiative, release 4; IEU Open GWAS Project identifier: ebi-a-GCST010780; https://gwas.mrcieu.ac.uk/datasets/ 11.Covid-19 (round 6): Covid-19 Host Genetics Initiative, release 6: https://www.covid19hg.org/results/r6/ 12.Job involves heavy, manual labor or physical work: MRC-IEU; IEU Open GWAS Project identifier: ukb-b-2002; https://gwas.mrcieu.ac.uk/datasets/ 13.Noisy workplace: MRC-IEU; IEU Open GWAS Project identifier: ukb-b-2091; https://gwas.mrcieu.ac.uk/datasets/ 14.Antisocial behavior (ASB) Broad Antisocial Behavior Consortium (BroadABC); http://broadabc.ctglab.nl/ (Newest release under review: summary data available upon request via) 15.Townsend Deprivation Index: MRC-IEU; IEU Open GWAS Project identifier: ukb-b-10011; https://gwas.mrcieu.ac.uk/datasets/ 16.Gastrointestinal diseases: FINNGen Biobank analysis; 39,639 cases and 56,860 controls (European); binary; IEU Open GWAS Project identifier: finn-a-K11_GIDISEASES; https://www.finngen.fi/fi/ 17.Chronic obstructive pulmonary disease: UKBB/Neale lab; IEU Open GWAS Project identifier: ukb-d-COPD_EXCL; https://gwas.mrcieu.ac.uk/datasets/ 18.Genitourinary diseases: UKBB/Neale lab; IEU Open GWAS Project identifier: ukb-d-XIV_GENITOURINARY; https://gwas.mrcieu.ac.uk/datasets/ 19.Neuroticism score: MRC-IEU; IEU Open GWAS Project identifier: ukb-b-4630; https://gwas.mrcieu.ac.uk/datasets/ 20.Seen doctor for nerves, anxiety, tension, or depression: MRC-IEU; IEU Open GWAS Project identifier: ukb-b-6991; https://gwas.mrcieu.ac.uk/datasets/ 21.Plays computer games: MRC-IEU; IEU Open GWAS Project identifier: ukb-b-4779; https://gwas.mrcieu.ac.uk/datasets/ 22.Victim of physically violent crime: UKBB/Neale lab; IEU Open GWAS Project identifier: ukb-d-20529; https://gwas.mrcieu.ac.uk/datasets/ 23.Risk tolerance: Karlsson Linner et al. (2019); Social Science Genetic Association Consortium (SSGAC); https://www.thessgac.org/ 24.Witnessed sudden violent death: UKBB/Neale lab; IEU Open GWAS Project identifier: ukb-d-20530; https://gwas.mrcieu.ac.uk/datasets/

### Summary of Updates

This version of the manuscript has been revised to update the following: Abstract and presentation of findings.

